# Evaluating the inclusion of lesbian, gay, bisexual, transgender, and queer-related content in graduate medical education: a national survey of program directors

**DOI:** 10.1101/2025.02.15.25322332

**Authors:** Andrew M. Pregnall, Joshua A. Pesantes, Gennady Vulakh, Youvin Chung, Setareh Gooshvar, C. Jessica Dine

## Abstract

**Background:** National organizations have identified incorporation of LGBTQ-health content into graduate medical education programs as key action items; however, there has been no systematic study of LGBTQ-health content in these programs.

**Objective:** The primary objective of this study was to systematically evaluate the quantity of LGBTQ-related didactic and clinical education in graduate medical education programs.

**Methods:** A cross-sectional, internet-based survey study of programs participating in the 2023-2024 Electronic Residency Application System, performed from September 2023-August 2024.

**Results:** Of 4,512 programs, 1,048 programs responded (23.2%). The median and mean number of didactic hours per year dedicated to LGBTQ-related content was 2.0 (IQR, 1.0–5.0, range, 0.0-200.0) and 4.0 (SD 9.1), respectively. The median and mean number of clinical hours per year dedicated to LGBTQ-related content was 10.0 (IQR, 1.5 – 40.0, range, 0.0 – 2000.0) and 61.0 (SD 188.4), respectively. Multiple programs reported that residents received no exposure to LGBTQ-related health content in either didactic settings (15.8%; 95% CI, 13.5%-18.5%) or clinical settings (19.4%, CI 16.1% – 23.0%). The most covered didactic topics were gender identity (43.6%), sexual orientation (41.6%), and barriers to care (32.0%). The most covered clinical topics were Pre-Exposure Prophylaxis/Post-Exposure Prophylaxis (77.0%) and facial masculinization/feminization surgery (68.4%). The most cited barriers to including LGBTQ-related health topics were the lack of faculty with requisite knowledge/expertise (56.1%, CI 52.3% – 59.9%) and the lack of time (48.3%, CI 44.5% – 52.1%).

**Conclusions:** Multiple programs provide no didactic or clinical exposure to LGBTQ-related health topics, which does not align with the goals outlined by national organizations.

## Background

Lesbian, gay, bisexual, transgender, queer, gender-nonconforming, and those born with differences in sexual development — collectively referred to as LGBTQ people or sexual and gender minorities (SGM) — composed 7.6% of the US population and 22.3% of Generation Z (those born between 1997 and 2012) in 2023.^1^ Despite their prevalence, LGBTQ people are known to face many societal and health disparities, underscoring the need for competent and affirming medical care.^2–5^ Although studies have examined LGBTQ-related education within individual medical specialties,^6–11^ no study has systematically explored LGBTQ-related content across graduate medical education (GME) programs.

In 2011, medical schools dedicated a median of 5 hours to LGBTQ content, with over one-third of schools reporting that they dedicated 0 hours.^12^ Since then, several shifts have occurred within American society and within medical care concerning LGBTQ issues. For example, the Association of American Medical Colleges (AAMC) released a 2014 report on incorporating LGBTQ topics into undergraduate medical education (UME), and a 2020 National Academies report highlighted the need for LGBTQ-related cultural and clinical competency training for clinicians.^4,5^ A 2024 follow-up study on UME found that the average curricular hours increased, from 5 to 11 hours per year, but highlighted that more changes are needed to provide quality care to LGBTQ people and to combat existing disparities.^13^

At the GME level, several studies have detailed the inclusion of LGBTQ-related content in specific medical specialties. Surveys of emergency medicine program directors from 2014 and 2021 demonstrated an increase in the proportion of programs including LGBTQ-related didactic content from 26% to 75% as well as a rise in the median hours of teaching from 0 to 2 per year.^6,9^ A 2017 study of urology and plastic surgery programs reported median didactic times of 1 and 2 hour(s) spent on transgender health, respectively. Additionally, while 56% of program directors indicated that transgender health was an important topic, others indicated they would not change their curricula until mandated by accrediting organizations.^7^ Similarly, a 2024 survey found that 80% of dermatology program directors reported SGM health topics as important to include in their curricula; however, 46% of programs had 0 hours dedicated to SGM content.^11^ Lastly, a recent survey of pediatrics program directors reported that 54% of respondents believed their residents were either not at all, or only somewhat, prepared to care for LGBTQ patients after training.^10^

Given the lack of comprehensive data on the inclusion of LGBTQ-related content in GME programs, this study thus aims to systematically evaluate the quantity of LGBTQ-related didactic and clinical education through a national survey of program directors. It secondarily aims to describe the topics covered in didactic and clinical education as well as existing barriers and strategies for success faced/utilized by programs.

## Methods

### Study design

We performed a cross-sectional internet-based survey of program directors and program coordinators of residency programs participating in the 2023-2024 Electronic Residency Application System or Plastic Surgery Common Application. The University of Pennsylvania Institutional Review Board deemed this study exempt.

### Survey development and distribution

Two authors (AMP and CJD) developed a survey using questions from surveys with evidence of strong validity as a template, modifying them to reflect changes in the field of LGBTQ health.^6,7,9,12^ After development, we provided the draft survey to two associate program directors with expertise in LGBTQ health and revised the survey based on their feedback. An annotated version of the survey with citations to the original studies, our changes to the questions, and associated reasoning is provided in **Supplementary Appendix 1**.

We distributed survey invitations via email from September 2023 to August 2024. We sent monthly email reminders to programs that had not started the survey or who had incomplete surveys. We administered the survey through REDCap (Vanderbilt University; Nashville, TN, USA) using a unique identifier and link for each program to avoid data duplication. Each unique link first directed participants to an informed consent page. Participating program directors were allowed to designate other individuals to complete the survey by utilizing a unique link and return code to forward the survey to designated individuals. We collected no information about the individuals completing the survey and kept all program-level responses confidential.

### Study outcomes

The primary outcome of our survey was the number of didactic hours GME programs dedicated to teaching LGBTQ-related topics per year. Secondary outcomes of our study reported here include (1) the number of clinical hours per year dedicated to the clinical care of LGBTQ patients; (2) topics covered in didactic teaching and clinical experiences; (3) barriers to integrating LGBTQ-related topics into GME programs; and (4) strategies that have been, or are expected to be, successful in increasing LGBTQ-related topic coverage in GME programs.

### Statistical analysis

We utilized an available case analysis to include all available responses for each survey question. First, we utilized the Chi-square test to compare responders versus non-responders. We then calculated descriptive statistics for all survey questions. Given non-normal distributions of the number of clinical and didactic hours, we utilized the Kruskal-Wallis test to compare hours of teaching by medical specialty, program type, and region and to examine variation within medical specialties by program type and region. We used the Fisher exact test to examine (1) differences in programs reporting zero versus non-zero didactic/clinical hours and (2) barriers to incorporating LGBTQ-related content by medical specialty. We utilized the Chi-square test to examine these topics by program type and region. We performed all analyses using R Version 4.2.1.

## Results

### Characterization of respondents and nonrespondents

Of 4,512 programs, we received 1,048 responses (23.2%). Of these, 62.6% fully completed the survey (n = 656), 28.1% partially completed the survey (n = 294), and 9.4% declined to participate (n = 98). The response rate by specialty, geographic region, and program type is presented in **Table 1**. There was a statistically significant difference in responders versus non-responders based on medical specialty (P < 0.001, Chi-square test) and program type (P = 0.002, Chi-square test). There was no difference based on geographic region (P = 0.173, Chi-square test). Finally, we conducted a sensitivity analysis of our findings by limiting data analysis to programs with complete survey responses. The results varied by less than 5% from those reported below.

**Table 1:** Characterization of Survey Respondents and Non-Respondents, Stratified by Specialty, Region, and Program Type.

|  | **Responded<sup>a</sup>**<br>N (%) | **Not Responded<sup>b</sup>**<br>N (%) | **P-Value<sup>c</sup>** | **Response**<br>**Rate (%)** |
| --- | --- | --- | --- | --- |
|  | 950 | 3562 |  |  |
| **Specialty** |  |  |  |  |
| Anesthesiology | 51 (5.4) | 96 (2.7) |  | 34.0 |
| Child Neurology | 12 (1.3) | 55 (1.5) |  | 17.9 |
| Dermatology | 19 (2.0) | 107 (3.0) |  | 15.1 |
| Diagnostic Radiology | 41 (4.3) | 135 (3.8) |  | 23.3 |
| Emergency Medicine | 79 (8.3) | 193 (5.4) |  | 29.0 |
| Family Medicine | 129 (13.6) | 521 (14.6) |  | 19.7 |
| Internal Medicine | 92 (9.7) | 497 (14.0) |  | 15.6 |
| Interventional Radiology | 9 (0.9) | 79 (2.2) |  | 10.2 |
| Neurological Surgery | 21 (2.2) | 78 (2.2) |  | 21.2 |
| Neurology | 47 (4.9) | 108 (3.0) |  | 30.3 |
| Obstetrics and Gynecology | 61 (6.4) | 201 (5.6) |  | 22.9 |
| Orthopedic Surgery | 31 (3.3) | 142 (4.0) | <0.001 | 17.3 |
| Otolaryngology | 19 (2.0) | 103 (2.9) |  | 15.6 |
| Pathology | 26 (2.7) | 100 (2.8) |  | 20.6 |
| Pediatrics | 71 (7.5) | 151 (4.2) |  | 32.0 |
| Physical Medicine and<br>Rehabilitation | 24 (2.5) | 73 (2.0) |  | 24.7 |
| Plastic Surgery | 7 (0.7) | 84 (2.4) |  | 7.7 |
| Psychiatry | 59 (6.2) | 210 (5.9) |  | 21.6 |
| Public Health | 10 (1.1) | 18 (0.5) |  | 35.7 |
| Radiation Oncology | 12 (1.3) | 55 (1.5) |  | 17.9 |
| Surgery | 60 (6.3) | 243 (6.8) |  | 19.5 |
| Transition Year | 25 (2.6) | 113 (3.2) |  | 18.2 |
| Urology | 24 (2.5) | 99 (2.8) |  | 19.5 |
| Vascular Surgery | 10 (1.1) | 49 (1.4) |  | 17.0 |
| Other | 11 (1.2) | 52 (1.5) |  | 17.7 |
| **Region** |  |  |  |  |
| Midwest | 235 (24.7) | 864 (24.3) |  | 26.4 |
| Northeast | 251 (26.4) | 914 (25.7) | 0.173 | 21.5 |
| South | 293 (30.8) | 1217 (34.2) |  | 19.4 |
| West | 171 (18.0) | 562 (15.8) |  | 23.3 |
| **Program Type** |  |  |  |  |
| Community-based | 136 (14.3) | 603 (16.9) |  | 18.4 |
| Community-based university affiliated | 325 (34.2) | 1381 (38.8) | 0.002 | 19.0 |
| Military-based | 0 (0.0) | 3 (0.1) |  | 0.0 |
| University-based | 476 (50.1) | 1537 (43.1) |  | 23.6 |
| Other | 13 (1.4) | 38 (1.1) |  | 25.5 |

<sup>a</sup>Responded indicates both partial and complete responses to the survey.
<sup>b</sup>Not responded includes both programs that did not respond and those that declined to respond.
<sup>c</sup>The p-values for specialty, region, and program type are the result of the Chi-square test.

### Hours of LGBTQ-related didactic content and clinical exposure in GME programs

We received 871 and 527 responses to our questions on the number of didactic and clinical hours GME programs dedicate to LGBTQ-related topics, respectively. The median number of didactic hours per year across all programs was 2.0 (IQR, 1.0–5.0, range, 0.0-200.0). The corresponding mean (SD) was 4.0 (9.1). The number of didactic hours stratified by specialty, region, and program type are presented in **Table 2**. There was a statistically significant difference in the number of didactic hours based on specialty (P < 0.001; Kruskal-Wallis test). Family medicine programs reported the greatest average didactic hours (mean [median]: 7.7 [4.0]) with internal medicine programs (6.2 [4.0]) and pediatrics programs (6.0 [4.0]) reporting the second and third most hours, respectively. Conversely, interventional radiology (1.0 [0.0]), pathology (0.9 [0.5]), and neurosurgery programs (1.4 [0.5]) reported the least coverage. There was also a statistically significant difference in the number of didactic hours based on program type (P = 0.005; Kruskal-Wallis test). Community-based university-affiliated programs dedicated the most time to LGBTQ topics (5.0 [3.0]) while university-based programs dedicated the least time (3.5 [2.0]). There was no difference in the number of didactic hours based on region. Within medical specialties, there was no statistically significant difference in the number of didactic hours based on region; however, there were differences based on program type within anesthesia, family medicine, and transition year programs (see **Supplementary Table 1**).

**Table 2:** Descriptive Statistics for the Number of Didactic and Clinical Hours Dedicated to LGBTQ Content in Graduate Medical Education, Stratified by Specialty, Region, and Program Type.

|  | Didactic Hours |  |  |  | Clinical Hours |  |  |  |
| --- | --- | --- | --- | --- | --- | --- | --- | --- |
|  | Median (IQR) | Range | Mean (SD) | P-Value<sup>a</sup> | Median (IQR) | Range | Mean (SD) | P-Value<sup>a</sup> |
| Specialty (n, n)<sup>b</sup> |  |  |  |  |  |  |  |  |
| Anesthesiology (48, 25) | 2.0 (0.0-2.3) | 0.0 – 20.0 | 2.4 (3.4) |  | 20.0 (2.0-100.0) | 0.0 – 2000.0 | 157.4 (432.1) |  |
| Child Neurology (12, 6) | 1.5 (0.0-2.0) | 0.0 – 4.0 | 1.3 (1.2) |  | 8.5 (0.5-15.0) | 0.0 – 20.0 | 8.7 (9.0) |  |
| Dermatology (18, 8) | 2.5 (2.0-4.0) | 1.0 – 12.0 | 3.8 (3.2) |  | 11.0 (7.5-70.0) | 0.0 – 100.0 | 36.5 (43.6) |  |
| Diagnostic Radiology (35, 19) | 1.0 (0.5-2.0) | 0.0 – 12.0 | 1.8 (2.2) |  | 0.0 (0.0-1.0) | 0.0 – 10.0 | 1.5 (3.2) |  |
| Emergency Medicine (71, 26) | 3.0 (2.0-4.5) | 0.0 – 10.0 | 3.3 (2.2) |  | 9.0 (2.0-24.8) | 0.0 – 365.0 | 31.3 (73.2) |  |
| Family Medicine (120, 91) | 4.0 (3.0-8.0) | 0.0 – 200.0 | 7.7 (18.4) |  | 16.0 (6.0-50.0) | 0.0 – 1000.0 | 73.2 (161.9) |  |
| Internal Medicine (84, 58) | 4.0 (2.0-5.3) | 0.0 – 120.0 | 6.2 (13.7) |  | 10.0 (3.0-28.9) | 0.0 – 960.0 | 42.9 (129.9) |  |
| Interventional Radiology (7, 3) | 0.0 (0.0-1.5) | 0.0 – 4.0 | 1.0 (1.5) |  | – | – | – |  |
| Neurological Surgery (20, 11) | 0.5 (0.0-2.0) | 0.0 – 10.0 | 1.4 (2.4) |  | 1.0 (0.0-40.0) | 0.0 – 1000.0 | 103.4 (298.0) |  |
| Neurology (44, 22) | 1.5 (1.0-3.0) | 0.0 – 150.0 | 2.3 (2.7) |  | 5.0 (1.0-10.0) | 0.0 – 200.0 | 21.9 (46.1) |  |
| Obstetrics and Gynecology (59, 42) | 4.0 (2.0-6.0) | 0.0 – 20.0 | 4.7 (3.6) |  | 20.0 (6.5-50.0) | 0.0 – 1500.0 | 94.2 (270.9) |  |
| Orthopedic Surgery (27, 15) | 1.0 (0.0-2.0) | 0.0 – 5.0 | 1.3 (1.6) | < 0.001 | 1.0 (0.0-20.0) | 0.0 – 500.0 | 46.9 (128.3) | < 0.001 |
| Otolaryngology (18, 15) | 2.5 (0.3-4.0) | 0.0 – 10.0 | 2.8 (2.7) |  | 10.0 (6.0-50.0) | 1.0 – 100.0 | 30.2 (35.9) |  |
| Pathology (23, 18) | 0.5 (0.0-2.0) | 0.0 – 4.0 | 0.9 (1.2) |  | 0.0 (0.0-0.0) | 0.0 – 100.0 | 6.4 (23.5) |  |
| Pediatrics (61, 41) | 4.0 (2.0-6.0) | 0.0 – 80.0 | 6.0 (10.4) |  | 15.0 (6.0-40.0) | 0.0 – 300.0 | 37.8 (60.2) |  |
| Physical Medicine and Rehabilitation (23, 13) | 1.0 (0.0-2.0) | 0.0 – 8.0 | 1.2 (1.8) |  | 0.0 (0.0-3.0) | 0.0–200.0 | 16.5 (55.2) |  |
| Plastic Surgery (6, 4) | 3.5 (2.3-4.8) | 2.0 – 14.0 | 5.0 (4.6) |  | – | – | – |  |
| Psychiatry (55, 31) | 4.0 (2.0-5.0) | 0.0 – 30.0 | 4.7 (4.8) |  | 10.0 (2.5-80.0) | 0.0 – 1440.0 | 152.1 (350.5) |  |
| Public Health (9, 5) | 4.0 (2.0-5.0) | 0.0 – 10.0 | 3.8 (3.1) |  | 10.0 (10.0-30.0) | 0.0 – 50.0 | 20.0 (20.0) |  |
| Radiation Oncology (12, 7)) | 1.5 (0.8-2.3) | 0.0 – 5.0 | 1.7 (1.5) |  | 2.0 (2.0 – 15.0) | 0.0 – 30.0 | 9.4 (12.5) |  |
| Surgery (54, 26) | 1.5 (0.1-3.0) | 0.0-12.0 | 2.2 (2.6) |  | 4.0 (0.1 – 17.5) | 0.0 – 1000.0 | 55.0 (196.8) |  |
| Transition Year (24, 12) | 2.5 (1.8-5.0) | 0.0 – 36.0 | 4.7 (7.3) |  | 22.5 (1.5 – 37.5) | 0.0 – 100.0 | 29.8 (33.5) |  |
| Urology (23, 19) | 3.0 (1.0-4.0) | 0.0 – 10.0 | 3.0 (2.6) |  | 15.0 (8.0 – 65.0) | 0.0 – 600.0 | 83.9 (168.2) |  |
| Vascular Surgery (8, 2) | 1.0 (0.0-1.3) | 0.0 – 2.0 | 0.9 (0.8) |  | – | – | – |  |
| Other (10, 19) | 1.0 (0.0-2.0) | 0.0 – 12.0 | 2.2 (3.7) |  | 2.0 (0.0-60.0) | 0.0 – 500.0 | 55.5 (121.0) |  |
| Region (n , n)<sup>b</sup> |  |  |  |  |  |  |  |  |
| Midwest (211, 133) | 2.0 (1.0-4.0) | 0.0 – 80.0 | 4.0 (6.9) |  | 10.0 (2.0-40.0) | 0.0 – 1000.0 | 50.8 (135.7) |  |
| Northeast (224, 130) | 3.0 (1.0-5.0) | 0.0 – 120.0 | 4.3 (8.7) |  | 10.0 (2.0-25.0) | 0.0 – 2000.0 | 54.8 (213.6) |  |
| South (275, 157) | 2.0 (1.0-4.0) | 0.0 – 40.0 | 3.3 (4.2) | 0.101 | 10.0 (1.0-40.0) | 0.0 – 1000.0 | 46.1 (129.7) | 0.479 |
| West (161, 107) | 3.0 (1.0-5.0) | 0.0 – 200.0 | 5.1 (15.9) |  | 10.0 (1.0-50.0) | 0.0 – 1500.0 | 103.0 (265.8) |  |
| **Program Type (n, n)<sup>b</sup>** |  |  |  |  |  |  |  |  |
| Community-based (126, 70) | 3.0 (1.0-5.0) | 0.0 – 36.0 | 3.8 (4.6) |  | 10.0 (1.3-49.5) | 0.0 – 625.0 | 44.9 (95.6) |  |
| Community-based university affiliated (299, 206) | 3.0 (1.0-5.0) | 0.0 – 200.0 | 5.0 (13.7) |  | 10.0 (2.0-37.5) | 0.0 – 1000.0 | 49.5 (134.5) |  |
| Military-based (0, 0) | – | – | – | 0.005 | – | – | – | 0.668 |
| University-based (434, 246) | 2.0 (1.0-4.0) | 0.0 – 80.0 | 3.5 (5.6) |  | 10.0 (1.0-40.0) | 0.0 – 2000.0 | 72.2 (233.5) |  |
| Other (12, 5) | 2.5 (0.4-5.0) | 0.0 – 12.0 | 3.6 (4.2) |  | 20.0 (3.0-20.0) | 0.0 – 1000.0 | 208.6 (442.5) |  |

<sup>a</sup>P-values are the result of the Kruskal-Wallis test.
<sup>b</sup>N, n represents the total number of programs with available data to estimate the didactic hours and clinical hours, respectively

One hundred thirty-eight programs (15.8%; 95% CI, 13.5%-18.5%) reported 0 hours of LGBTQ-related didactic content (see **Supplementary Table 2**). Programs reporting 0 hours of coverage varied significantly by specialty and program type (P < 0.001 and P = 0.023, respectively; Chi-square test). Dermatology, plastic surgery, and emergency medicine had the lowest proportion of programs that dedicated no time towards LGBTQ didactic content (0.0% (n = 0), 0.0% (n = 0), and 1.4% (n = 1), respectively) while interventional radiology, neurosurgery, and pathology had the highest proportion of programs (57.1% (n = 4), 50.0% (n = 10), and 47.8% (n = 11), respectively). Meanwhile, 19.1% of university-based programs (n = 83) dedicated no time towards LGBTQ didactic content compared to 11.0% of community-based university affiliated programs (n = 33).

The median number of clinical hours dedicated per year was 10.0 (IQR, 1.5 – 40.0, range, 0.0 – 2000.0). The corresponding mean (SD) was 61.0 (188.4). The number of clinical hours stratified by specialty, region, and program type is presented in **Table 2**. There was a statistically significant difference in the number of clinical hours based on specialty (P < 0.001; Kruskal-Wallis test) but not by region or program type. Transition year programs reported the highest median clinical hours at 22.5 hours (IQR: 1.5 – 37.5) with anesthesiology (20.0 [2.0-100.0]) and obstetrics/gynecology (20.0 [6.5-50.0]) programs reporting the next greatest amounts. Conversely, pathology (0.0 [0.0-0.0]), diagnostic radiology (0.0 [0.0-1.0]), and physical medicine and rehabilitation (0.0 [0.0-3.0]) programs reported the least exposure. Within medical specialties there was no difference in the number of clinical hours based on region; however, there were possible differences in the number of clinical hours based on program type in psychiatry (P = 0.037, Kruskal-Wallis test), transition year (P = 0.038), anesthesiology (P = 0.042), OBGYN (P = 0.046), and dermatology (P = 0.079) programs (see **Supplementary Table 3**). One hundred and two programs dedicated 0 hours to clinical coverage of LGBTQ patients (19.4%, CI 16.1% – 23.0%). Programs reporting 0 hours of coverage varied significantly by specialty and (P = 0.046, Fisher exact test) and possibly varied by program type (P = 0.061, Chi-square test); see **Supplementary Table 2**). Otolaryngology, pediatrics, and family medicine had the lowest proportion of programs that dedicated no time towards LGBTQ clinical content (0.0% (n = 0), 2.4% (n = 1), and 7.7% (n = 7), respectively) while pathology, diagnostic radiology, and physical medicine and rehabilitation had the highest proportion of programs (77.8% (n = 14), 66.4% (n = 13), and 53.8% (n = 7), respectively). Meanwhile, 23.6% of university-based programs (n = 58) dedicated no time towards LGBTQ clinical content compared to 13.6% of community-based university affiliated programs (n = 28).

### Topics covered in LGBTQ-related didactics and clinical experiences

We received between 762 and 768 responses on LGBTQ-related topics covered in didactic education. The most common topics covered by programs each year included gender identity at 334 programs (43.6%; CI, 40.1% – 47.1%), sexual orientation at 319 programs (41.6%; CI, 38.2% – 45.1%), and barriers to care at 246 programs (32.0%; CI, 28.7% – 35.3%) (see **Table 3**). The least common topics were elder health at 458 programs (60.0%; CI, 56.6% – 63.5%), coming out at 375 programs (49.0%; CI, 45.4% – 52.5%), and LGBTQ people of color at 370 programs (48.6%; CI, 45.0% – 52.1%).

**Table 3:**
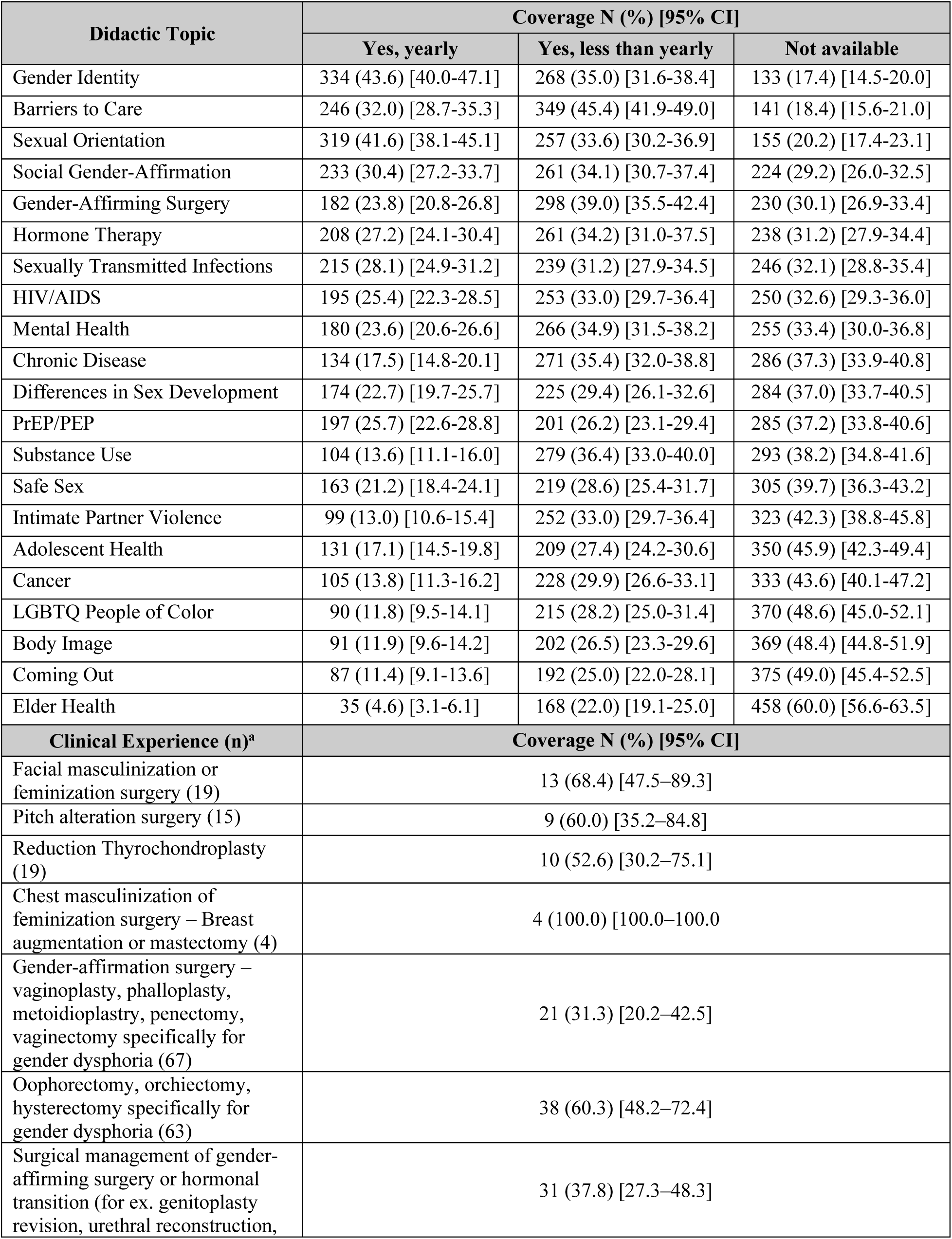
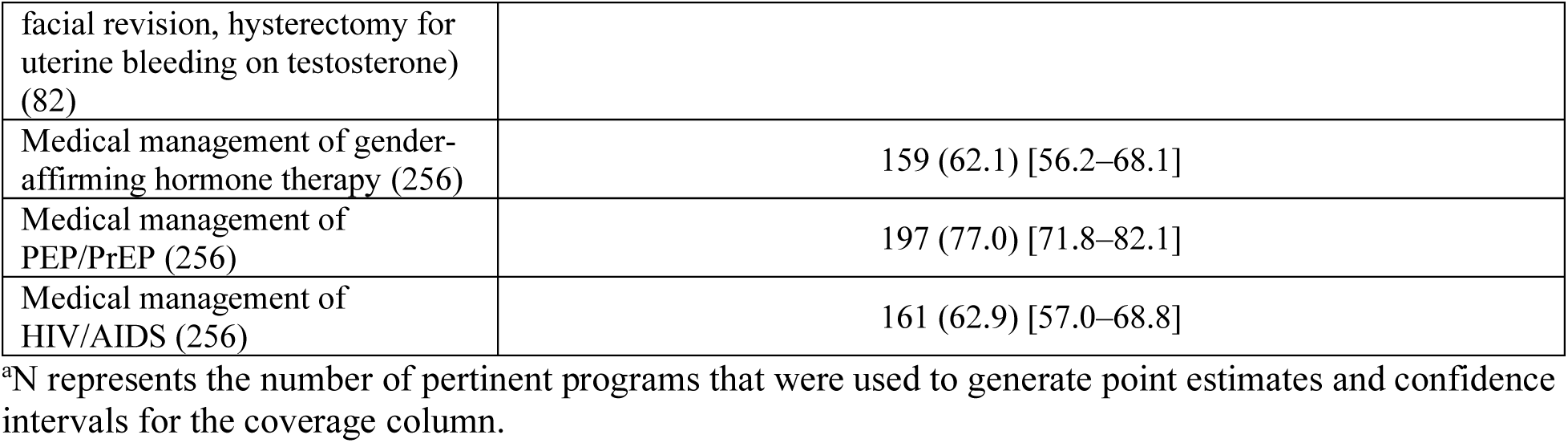
Descriptive Statistics for LGBTQ-Health Related Didactic Topics and Clinical Experiences in Graduate Medical Education.

| Didactic Topic | Coverage N (%) [95% CI] |  |  |
| --- | --- | --- | --- |
|  | Yes, yearly | Yes, less than yearly | Not available |
| Gender Identity | 334 (43.6) [40.0-47.1] | 268 (35.0) [31.6-38.4] | 133 (17.4) [14.5-20.0] |
| Barriers to Care | 246 (32.0) [28.7-35.3] | 349 (45.4) [41.9-49.0] | 141 (18.4) [15.6-21.0] |
| Sexual Orientation | 319 (41.6) [38.1-45.1] | 257 (33.6) [30.2-36.9] | 155 (20.2) [17.4-23.1] |
| Social Gender-Affirmation | 233 (30.4) [27.2-33.7] | 261 (34.1) [30.7-37.4] | 224 (29.2) [26.0-32.5] |
| Gender-Affirming Surgery | 182 (23.8) [20.8-26.8] | 298 (39.0) [35.5-42.4] | 230 (30.1) [26.9-33.4] |
| Hormone Therapy | 208 (27.2) [24.1-30.4] | 261 (34.2) [31.0-37.5] | 238 (31.2) [27.9-34.4] |
| Sexually Transmitted Infections | 215 (28.1) [24.9-31.2] | 239 (31.2) [27.9-34.5] | 246 (32.1) [28.8-35.4] |
| HIV/AIDS | 195 (25.4) [22.3-28.5] | 253 (33.0) [29.7-36.4] | 250 (32.6) [29.3-36.0] |
| Mental Health | 180 (23.6) [20.6-26.6] | 266 (34.9) [31.5-38.2] | 255 (33.4) [30.0-36.8] |
| Chronic Disease | 134 (17.5) [14.8-20.1] | 271 (35.4) [32.0-38.8] | 286 (37.3) [33.9-40.8] |
| Differences in Sex Development | 174 (22.7) [19.7-25.7] | 225 (29.4) [26.1-32.6] | 284 (37.0) [33.7-40.5] |
| PrEP/PEP | 197 (25.7) [22.6-28.8] | 201 (26.2) [23.1-29.4] | 285 (37.2) [33.8-40.6] |
| Substance Use | 104 (13.6) [11.1-16.0] | 279 (36.4) [33.0-40.0] | 293 (38.2) [34.8-41.6] |
| Safe Sex | 163 (21.2) [18.4-24.1] | 219 (28.6) [25.4-31.7] | 305 (39.7) [36.3-43.2] |
| Intimate Partner Violence | 99 (13.0) [10.6-15.4] | 252 (33.0) [29.7-36.4] | 323 (42.3) [38.8-45.8] |
| Adolescent Health | 131 (17.1) [14.5-19.8] | 209 (27.4) [24.2-30.6] | 350 (45.9) [42.3-49.4] |
| Cancer | 105 (13.8) [11.3-16.2] | 228 (29.9) [26.6-33.1] | 333 (43.6) [40.1-47.2] |
| LGBTQ People of Color | 90 (11.8) [9.5-14.1] | 215 (28.2) [25.0-31.4] | 370 (48.6) [45.0-52.1] |
| Body Image | 91 (11.9) [9.6-14.2] | 202 (26.5) [23.3-29.6] | 369 (48.4) [44.8-51.9] |
| Coming Out | 87 (11.4) [9.1-13.6] | 192 (25.0) [22.0-28.1] | 375 (49.0) [45.4-52.5] |
| Elder Health | 35 (4.6) [3.1-6.1] | 168 (22.0) [19.1-25.0] | 458 (60.0) [56.6-63.5] |
| Clinical Experience (n)<sup>a</sup> | Coverage N (%) [95% CI] |  |  |
| Facial masculinization or feminization surgery (19) | 13 (68.4) [47.5–89.3] |  |  |
| Pitch alteration surgery (15) | 9 (60.0) [35.2–84.8] |  |  |
| Reduction Thyrochondroplasty (19) | 10 (52.6) [30.2–75.1] |  |  |
| Chest masculinization of feminization surgery – Breast augmentation or mastectomy (4) | 4 (100.0) [100.0–100.0] |  |  |
| Gender-affirmation surgery – vaginoplasty, phalloplasty, metoidioplasty, penectomy, vaginectomy specifically for gender dysphoria (67) | 21 (31.3) [20.2–42.5] |  |  |
| Oophorectomy, orchiectomy, hysterectomy specifically for gender dysphoria (63) | 38 (60.3) [48.2–72.4] |  |  |
| Surgical management of gender-affirming surgery or hormonal transition (for ex. genitoplasty revision, urethral reconstruction, | 31 (37.8) [27.3–48.3] |  |  |

|  |  |
| --- | --- |
| facial revision, hysterectomy for uterine bleeding on testosterone) (82) |  |
| Medical management of gender-affirming hormone therapy (256) | 159 (62.1) [56.2–68.1] |
| Medical management of PEP/PrEP (256) | 197 (77.0) [71.8–82.1] |
| Medical management of HIV/AIDS (256) | 161 (62.9) [57.0–68.8] |

<sup>a</sup>N represents the number of pertinent programs that were used to generate point estimates and confidence intervals for the coverage column.

We received 292 responses to our questions on the incorporation of clinical exposure to LGBTQ-related health topics amongst pertinent specialties (see **Table 3**). Between 23.0% and 68.7% of programs directors reported that residents received no exposure to LGBTQ-related clinical topics across multiple health domains. Additional topics covered in didactics and clinical experiences as reported by program directors in free text responses are available in **Supplementary Tables 4 and 5**.

### Barriers to incorporating LGBTQ-topics in GME and strategies for success

We received 656 responses to our questions on barriers to incorporating LGBTQ-content in GME and strategies for success (see **Table 4**). One hundred and forty-one programs reported no barriers to GME LGBTQ-content inclusion (21.5%, CI 18.4% – 24.6%). The most cited barriers were the lack of faculty with requisite knowledge/expertise at 368 programs (56.1%, CI 52.3% – 59.9%), and the lack of time at 317 programs (48.3%, CI 44.5% – 52.1%). Of note, 96 programs cited the influence of state and federal policies as a barrier to incorporating LGBTQ content into GME (14.6%, CI 11.9 – 17.3). The most common strategies programs cited as having been successful in increasing LGBTQ content were curricular materials focusing on LGBTQ health and health disparities (32.2%, CI 28.6 – 35.7) and faculty willing and able to teach on these topics (36.9%, CI 33.2 – 40.6). Several statistically significant differences in barriers were noted based on medical specialty, region, and program type (see **Supplementary Table 6**).

**Table 4:** Descriptive Statistics for Barriers to Including LGBTQ-Health Related Topics in Graduate Medical Education and Strategies for Success (n = 656)

| **Barriers** | **N (%) [95% CI]** |  |
| --- | --- | --- |
| Lack of interested faculty | 129 (19.7) [16.6 – 22.7] |  |
| Lack of faculty with requisite expertise/knowledge | 368 (56.1) [52.3 – 59.9] |  |
| Lack of funding | 98 (14.9) [12.2 – 17.8] |  |
| Lack of time | 317 (48.3) [44.5 – 52.1] |  |
| Perception education is not needed | 107 (16.3) [13.5 – 19.1] |  |
| Opposition to inclusion of LGBTQ health topics | 40 (6.1) [4.3 – 7.9] |  |
| Influence of state or federal policies and/or political environment | 96 (14.6) [11.9 – 17.3] |  |
| Lack of patient population | 90 (13.7) [11.1 – 16.4] |  |
| No barriers | 141 (21.5) [18.4 – 24.6] |  |
| **Strategies** | **Has been successful**<br>**N (%) [95% CI]** | **Would likely be successful**<br>**N (%) [95% CI]** |
| Curricular material focusing on LGBTQ-related health/health disparities | 211 (32.2) [28.6 – 35.7] | 331 (50.5) [46.6 – 54.3] |
| Faculty willing and able to teach LGBTQ-related curricular content | 242 (36.9) [33.2 – 40.6] | 311 (47.4) [43.6 – 51.2] |
| Increased financial resources | 46 (7.0) [5.1 – 9.0] | 337 (51.4) [47.5 – 51.2] |
| Logistical support for teaching LGBTQ-related curricular content | 71 (10.8) [8.4 – 13.2] | 387 (59.0) [55.2 – 62.8] |
| More time in the curriculum to be able to teach LGBTQ-related content | 87 (13.3) [10.7 – 15.9] | 405 (61.7) [58.0 – 65.5] |
| More evidence-based research regarding LGBTQ health/health disparities | 91 (13.9) [11.2 – 16.5] | 383 (58.4) [54.6 – 62.2] |
| More evidence-based research regarding the teaching of LGBTQ-related health topics | 75 (11.4) [9.0 – 13.9] | 391 (60.0) [55.8 – 63.4] |
| Curricular material coverage required by accreditation bodies | 72 (11.0) [8.6 – 13.4] | 381 (58.1) [54.3 – 61.9] |
| Questions based on LGBTQ health/health disparities onboard examinations | 60 (9.1) [6.9 – 11.4] | 395 (60.2) [56.5 – 64.0] |
| Methods to evaluate LGBTQ curricular content | 45 (6.9) [5.0 – 8.8] | 365 (55.6) [51.8 – 59.4] |

## Discussion

This study is the first to examine the state of LGBTQ-related content in GME programs by administering a unified survey across all medical specialties utilizing the Electronic Residency Application System. In doing so, we present data evaluating the inclusion of LGBTQ-related content for seventeen medical specialties for the first time, and we present updated and/or complementary data for seven specialties. The most important finding of this study is that an estimated 15.8% and 19.4% of programs provide no LGBTQ-related didactic and clinical education, respectively.

Currently, the enhancement and expansion of LGBTQ-related content in medical education at both the undergraduate and graduate levels have been identified as key action items in the advancement of inclusive care in the US.^4,5,14–16^ Our data demonstrates that — across all surveyed medical specialties — programs dedicated a median of 2.0 didactic hours to LGBTQ-related content and estimated that residents receive a median of 10.0 hours of clinical exposure to LGBTQ-related topics per year. While our data is descriptive and cannot answer the question of whether this coverage is adequate, other studies have examined this question. For example, when examining readiness to address LGBTQ health issues among 120 internal medicine residents prior to a didactic, Streed et al. found no significant difference in pre- and post-test performance among internal medicine residents, indicating a generally low baseline knowledge that did not improve with increasing post-graduate year.^17^ A similar 2024 study assessing knowledge on care for transgender and nonbinary patients found that current and recently graduated family medicine residents did not feel that their training was adequate to provide care for this patient population^18^

Small studies surveying residents on their preparedness to care for LGBTQ patients have reported similar findings in obstetrics/gynecology,^19^ dermatology,^20^ emergency medicine,^21^ psychiatry,^22^ urology,^23^ and pediatrics.^24^ Given these data, we posit that the current depth of coverage of LGBTQ health topics in GME and the methodology for teaching these topics is inadequate in preparing future providers to care for this vulnerable population.

We found that multiple programs dedicated 0 hours to the administration of LGBTQ-related didactic content (15.8%; 95% CI, 13.5%-18.5%). Across the surveyed domains of didactic education, between 6.3-18.5% of program directors rated coverage as “Not Needed”, which varied broadly across medical specialties. While not all domains of LGBTQ health are pertinent to all medical specialties, notable specialty-specific clinical and cultural competencies exist.^25^ We found instances where didactic education is perceived to be unnecessary in domains that are explicitly pertinent to given medical specialties. For example, 18.8% of otolaryngology program directors reported that didactic coverage of gender-affirming surgery was not needed, despite otolaryngologists providing gender-affirming surgeries in the form of facial feminization/masculinization, reduction thyrochondroplasties, and pitch alteration surgeries. We similarly found multiple programs estimated that residents received no clinical exposure to pertinent LGBTQ-related topics (19.4%, CI 16.1% – 23.0%). For example, 23.0% of family medicine, obstetrics/gynecology, internal medicine, and pediatrics program directors reported that residents received no clinical exposure to the management of Pre-Exposure Prophylaxis/Post-Exposure Prophylaxis, which are essential tools in reducing the rates of HIV/AIDS infections. This dearth of didactic and clinical coverage ultimately does not align with the goals identified by national organizations.

We found that 21.5% of program directors reported no barriers to incorporating LGBTQ-related content in their respective GME programs. However, 56.1% of program directors reported that a lack of faculty with requisite expertise/knowledge was a barrier to incorporating LGBTQ-related content. This finding underscores the importance of recruiting a diverse set of individuals into medical schools, residency programs, and faculty positions.^26^ Additionally, 48.3% of program directors reported that a lack of time was a barrier to incorporating LGBTQ-related content. Given historic concerns that current training structures are inadequate in preparing residents to care for patients^27,28^ and the need to prepare residents to care for other disadvantaged communities, this finding highlights the importance of thoughtfully considering how to provide residents with the necessary training to provide clinically excellent care to diverse sets of patients who are often impacted by multiple social determinants of health.^29–31^

Our study also has several limitations. The main limitation of our study is the response rate (23.2%). Our response rate may have been impacted by factors such as survey fatigue which we attempted to mitigate through once monthly contact, leadership changes, and institutional, state, or federal policy environments. Our study is also limited by the possibility of response bias. For example, programs with more favorable attitudes towards LGBTQ health may have been more inclined to participate in the survey, or program directors may have overestimated their inclusion of LGBTQ-health topics due to social desirability effects. During data collection, several program directors indicated that they would not participate in our survey or indicated that they do not teach LGBTQ-health topics given current legislation in their state. Within the context of these limitations, we posit our data may overestimate the true extent to which LGBTQ topics are included in GME programs while underestimating regional differences. Future directions for this research should focus on (1) assessment of quality of existing education and interventions to increase the quality and quantity of teaching on LGBTQ topics and (2) elucidating the relationship between state and federal policies and training hours in order to provide actionable insight for policymakers and medical educators to promote LGBTQ health education.

## Conclusion

Our study provides foundational data to medical educators and researchers interested in improving LGBTQ health and health outcomes through reforms to medical education. While we found broad coverage of LGBTQ-related health topics in GME programs, we also found that this coverage falls short of goals identified by national organizations and that opportunities for improvement and innovation remain.

## Declarations

### Author Contributions

AMP and CJD envisioned the study and the designed the survey. AMP, JAP, GV, YC, and SG contributed to data collection and statistical analysis. All authors contributed to drafting and revision of the manuscript. AMP and CJD supervised the work.

### Consent for Publication

All authors agree to posting of this preprint on medRxiv.

### Previous Presentations

Interim data on barriers to inclusion and strategies for success were presented at the National LGBTQ Health Conference in Atlanta, Georgia in July 2024. Data on surgical specialties were presented at the American College of Surgeons Clinical Congress in San Francisco, California in October 2024. Data on medical specialties were presented at the American College of Physicians Southeastern Region Poster Day in October 2024.

### Data Availability

The datasets used and/or analyzed during the current study are available from the corresponding author on reasonable request.

### Funding

None.

## Acknowledgments

None.

## Disclaimers

None.

## Notes

### Competing Interest Statement

The authors have declared no competing interest.

### Funding Statement

This study did not receive any funding.

### Author Declarations

The University of Pennsylvania Institutional Review Board waived ethical approval for this work.

