## Supplemental File 1 for "Evaluating the inclusion of lesbian, gay, bisexual, transgender, and queer-related content in graduate medical education: a national survey of program directors"

This document presents the supplementary information referenced in "Evaluating the inclusion of lesbian, gay, bisexual, transgender, and queer-related content in graduate medical education: a national survey of program directors." A table of contents is listed below.

Supplementary Table 3: Average Number of Clinical Hours Dedicated to LGBTQ Patient Care in Graduate Medical Education by Medical Specialty, Stratified by Region and Program Type 14

Supplementary Table 4: Free Text Responses for Additional Types of Didactic Education ....... 15

Supplementary Table 5: Free Text Responses for Additional Types of Clinical Education ........ 16

Supplementary Table 6: Proportion of Programs Citing Barriers to Incorporating LGBTQ-Content into Graduate Medical Education, Stratified by Specialty, Region, and Program Type. 17

References ........................................................................................................................ 20

**Supplementary Appendix 1: Informed Consent and Survey Instrument**

**Question 1.**

We are conducting a research study about the incorporation of LGBTQ-related topics in the spectrum of undergraduate, graduate, and continuing medical education. The following survey should take about 15-20 minutes to complete. Participation is voluntary. If you would prefer not to participate, do not complete the survey. There is no penalty ifyou choose not to join the research study. If you agree to participate, please complete the attached survey. You should ask the study team any questions you have related to participating before agreeing to join the study. Please contact the principal investigator Jessica Dine, MD or co-investigator Andrew Pregnall, MSc, MPhil with any questions. Your responses are confidential. A deidentified version of the response data will be created upon cessation of data collection and all identifiable records will be deleted. We ask that you try to answer all questions. Data collected inthis study may be stored and shared for future research in a de-identified fashion. Upon cessation of data collection, it would not be possible for future researchers to identify you. This can be done without again seeking your consent in the future, as permitted by law. This research has been reviewed by the University of Pennsylvania Institutional Review Board (IRB). If you have any questions about your rights as a human research participant at any time before, during or after participation, please contact the University of Pennsylvania IRB at (215) 898-2614 for assistance.

**Informed Consent**

| | |
|---|---|
| ◯ | I certify that I have read the informed consent form. I understand my rights and responsibilities as well as those of the investigators as they have been presented here, and I affirm my wish to participate in this research study. |
| ◯ | I do not wish to participate in this research study. |

**Notes:** This question was adapted from Obedin-Maliver et al.[1] and standard informed consent verbiage from the University of Pennsylvania.

### Survey Part 1: Institutional Demographics

**Question 2.**

**Please choose your institution type:**

| | |
|---|---|
| ◯ | University-based training program |
| ◯ | Community-based training program |
| ◯ | Other [fill in the blank-box] |

**Notes:** This question was adapted from Morrison et al.[2] However, due to variability in the 'Other' responses we received, we abstracted institution type from the American Medical Association's FREIDA database to perform our final analysis.

**Question 3.**

**Please indicate where your program is located.**

| Drop down box indicating all 50 states, the District of Columbia, Puerto Rico |
|---|

***Notes***: This question was adapted from Moll et al.'s survey of emergency medicine program directors and Morrison et al.'s survey of urology and plastic surgery program directors.[2,3] It is intended to ascertain whether there are regional differences in the amount of LGBTQ-related content taught in graduate medical education programs. The analysis was performed based on US Census Region.

### Survey Part 2: Didactic Teaching of LGBTQ-Related Content in All Medical Specialties

**Question 4.**

**Please indicate to the nearest whole number how many AVERAGE hours are dedicated PER YEAR to DIDACTIC teaching of LGBTQ-related content in your training program:**

| ◯ [INPUT – accepts whole numbers] hour(s) |
|---|

**Notes:** This question has been adapted from all validated survey instuments.[1-4]

**Question 5.**

**Does your program provide education for residents in the following content areas at any point in their training:**

| | **Yes, we consistently cover this topic** | **Yes, but we inconsistency have covered this topic** | Not available | Don't know |
|---|---|---|---|---|
| Barriers to accessing medical care for LGBTQ people | ◯ | ◯ | ◯ | ◯ |
| Alcohol, tobacco, and/or other substance use amongst LGBTQ people | ◯ | ◯ | ◯ | ◯ |

| | | | | |
|---|---|---|---|---|
| Safer sex for LGBTQ people | ○ | ○ | ○ | ○ |
| **PrEP and PEP use for LGBTQ people** | ○ | ○ | ○ | ○ |
| HIV/AIDS in LGBTQ people | ○ | ○ | ○ | ○ |
| Sexually transmitted infections not including HIV/AIDS in LGBTQ people | ○ | ○ | ○ | ○ |
| Chronic disease risk for LGBTQ populations | ○ | ○ | ○ | ○ |
| Sexual orientation | ○ | ○ | ○ | ○ |
| Gender identity | ○ | ○ | ○ | ○ |
| **Differences in sex development**; intersex | ○ | ○ | ○ | ○ |
| Coming out | ○ | ○ | ○ | ○ |
| **Social gender-affirmation (e.g., wearing clothes or using a name in alignment with one's gender identity; a common first step of gender-affirmation for transgender and/or gender nonconforming adolescents)** | ○ | ○ | ○ | ○ |
| **Gender-affirming hormone therapy** | ○ | ○ | ○ | ○ |
| **Gender-affirming surgery (top surgeries such as breast augmentation or mastectomy; bottom surgeries such as phalloplasty or vaginoplasty; facial** | ○ | ○ | ○ | ○ |

| | | | | |
|---|---|---|---|---|
| **masculinization or feminization surgeries; orchiectomy, oophorectomy, or hysterectomy for gender dysphoria; etcetera)** | | | | |
| LGBTQ adolescent health | ◯ | ◯ | ◯ | ◯ |
| **LGBTQ elder health** | ◯ | ◯ | ◯ | ◯ |
| **Health of LGBTQ people of color** | ◯ | ◯ | ◯ | ◯ |
| Mental health in LGBTQ people | ◯ | ◯ | ◯ | ◯ |
| Body image in LGBTQ people | ◯ | ◯ | ◯ | ◯ |
| Unhealthy relationships (e.g., intimate partner violence) amongst LGBTQ people | ◯ | ◯ | ◯ | ◯ |

**Notes:** We adapted this question from Obedin-Maliver et al.[1] We changed the answer options from the 'required curriculum' versus 'elective curriculum' verbiage of the prior survey to reflect this survey's focus on graduate medical education. We added questions on PrEP/PEP, LGBTQ elder health, and health of LGBTQ people of color to reflect these new/underappreciated aspects of LGBTQ health. We changed the 'Disorders of Sex Development' to 'Differences in Sex Development' to reflect new, preferred language. We split 'Transitioning' into social gender-affirmation, gender-affirming hormone therapy, and gender-affirming surgery to delineate between three asepcts of gender-affirmation that transgender-nonbinary individuals may or may not choose to pursue.

**Question 6.**

**Please describe your opinions of how the following content areas are covered:**

| | Coverage not needed | Too little coverage | Basic coverage | In-depth coverage | Too much coverage | Don't know |
|---|---|---|---|---|---|---|
| Barriers to accessing medical care for LGBTQ people | ◯ | ◯ | ◯ | ◯ | ◯ | ◯ |
| Alcohol, tobacco, and/or other substance | ◯ | ◯ | ◯ | ◯ | ◯ | ◯ |

| | | | | | | |
|---|---|---|---|---|---|---|
| use amongst LGBTQ people | | | | | | |
| Safer sex for LGBTQ people | ◯ | ◯ | ◯ | ◯ | ◯ | ◯ |
| **PrEP and PEP use for LGBTQ people** | ◯ | ◯ | ◯ | ◯ | ◯ | ◯ |
| HIV/AIDS in LGBTQ people | ◯ | ◯ | ◯ | ◯ | ◯ | ◯ |
| Sexually transmitted infections not including HIV/AIDS in LGBTQ people | ◯ | ◯ | ◯ | ◯ | ◯ | ◯ |
| Chronic disease risk for LGBTQ populations | ◯ | ◯ | ◯ | ◯ | ◯ | ◯ |
| Sexual orientation | ◯ | ◯ | ◯ | ◯ | ◯ | ◯ |
| Gender identity | ◯ | ◯ | ◯ | ◯ | ◯ | ◯ |
| **Differences in sex development**; intersex | ◯ | ◯ | ◯ | ◯ | ◯ | ◯ |
| Coming out | ◯ | ◯ | ◯ | ◯ | ◯ | ◯ |
| **Social gender-affirmation (e.g., wearing clothes or using a name in alignment with one's gender identity; a common first step of gender-affirmation for transgender and/or gender nonconforming adolescents)** | ◯ | ◯ | ◯ | ◯ | ◯ | ◯ |
| **Gender-affirming hormone therapy** | ◯ | ◯ | ◯ | ◯ | ◯ | ◯ |
| **Gender-affirming surgery (top surgeries such as** | ◯ | ◯ | ◯ | ◯ | ◯ | ◯ |

| | | | | | | |
|---|---|---|---|---|---|---|
| **breast augmentation or mastectomy; bottom surgeries such as phalloplasty or vaginoplasty; facial masculinization or feminization surgeries; orchiectomy, oophorectomy, or hysterectomy for gender dysphoria; etcetera)** | | | | | | |
| LGBTQ adolescent health | ◯ | ◯ | ◯ | ◯ | ◯ | ◯ |
| **LGBTQ elder health** | ◯ | ◯ | ◯ | ◯ | ◯ | ◯ |
| **Health of LGBTQ people of color** | ◯ | ◯ | ◯ | ◯ | ◯ | ◯ |
| Mental health in LGBTQ people | ◯ | ◯ | ◯ | ◯ | ◯ | ◯ |
| Body image in LGBTQ people | ◯ | ◯ | ◯ | ◯ | ◯ | ◯ |
| Unhealthy relationships (e.g., intimate partner violence) amongst LGBTQ people | ◯ | ◯ | ◯ | ◯ | ◯ | ◯ |

**Notes:** We adapted this question from Obedin-Maliver et al.[1] The changes to the content of this question are the same as **Question 5**.

**Question 7.**

**Please indicate to the nearest whole number how many AVERAGE hours PER YEAR are dedicated to the clinical care of LGBTQ patients in your training program:**

| |
|---|
| ◯ [INPUT – accepts whole numbers] hour(s) |

**Notes:** This question has been adapted from all validated survey instuments.[1-4] It has been changed to reflect its focus on the clinical care of LGBTQ patients as opposed to didactic education. It most directly reflects a question used by Morrison et al.[2]

### Survey Part 3: Sub-Questions on Clinical Training for Pediatrics, Internal Medicine, Family Medicine, Plastic Surgery, Urology, Otolaryngology, and Obstetrics and Gynecology Programs

**Question 8.**

**Which topics do residents get direct exposure to in clinical settings in your training program. Please note that not all topics listed may apply to your medical specialty.**

| | |
|---|---|
| ☐ | Facial masculinization or feminization surgery |
| ☐ | **Pitch alteration surgery** |
| ☐ | **Reduction thyrochondroplastly (thyroid cartilage shave)** |
| ☐ | Top surgery — breast augmentation or mastectomy |
| ☐ | Bottom surgery — vaginoplasty, phalloplasty, metoidioplasty, penectomy, or vaginectomy specifically for gender dysphoria |
| ☐ | Surgical castration — oophorectomy, orchiectomy, hysterectomy specifically for gender dysphoria |
| ☐ | Surgical management of complications of gender confirming surgery or hormonal transmission (for example, genitoplasty revision, urethral reconstruction, facial revision, hysterectomy for uterine bleeding on testosterone) |
| ☐ | **Medical management of gender-affirming hormone therapy** |
| ☐ | **Medical management of PrEP/PEP** |
| ☐ | **Medical management of HIV/AIDS** |
| ☐ | Other [free-text] |

**Notes:** We adapted this question from Morrison et al.[2] We changed the verbiage of the question to focus solely on topics residents gained direct clinical exposure to as opposed to exposure both in clinical and didactic settings. We added 'Pitch alteration surgery' and 'Reduction thyrochondroplasty' as answer options to reflect gender-affirming surgical procedures performed by otolaryngologists. We added the 'Medical management of…' options to reflect medical aspects of LGBTQ healthcare.

**Question 9.**

**Please describe your opinions of how the following content areas are covered in clinical settings:**

| | Not applicable | Too little coverage | Basic coverage | In-depth coverage | Too much coverage | Don't know |
|---|---|---|---|---|---|---|

| | | | | | | |
|---|---|---|---|---|---|---|
| Facial masculinization or feminization surgery | ○ | ○ | ○ | ○ | ○ | ○ |
| **Pitch alteration surgery** | ○ | ○ | ○ | ○ | ○ | ○ |
| **Reduction thyrochondroplasty** | ○ | ○ | ○ | ○ | ○ | ○ |
| Top surgery — breast augmentation or mastectomy | ○ | ○ | ○ | ○ | ○ | ○ |
| Bottom surgery — vaginoplasty, phalloplasty, metoidioplasty, penectomy, or vaginectomy specifically for gender dysphoria | ○ | ○ | ○ | ○ | ○ | ○ |
| Surgical castration — oophorectomy, orchiectomy, hysterectomy specifically for gender dysphoria | ○ | ○ | ○ | ○ | ○ | ○ |
| Surgical management of complications of gender confirming surgery or hormonal transmission (for example, genitoplasty revision, urethral reconstruction, facial revision, hysterectomy for uterine bleeding on testosterone) | ○ | ○ | ○ | ○ | ○ | ○ |
| **Medical management of gender-affirming hormone therapy** | ○ | ○ | ○ | ○ | ○ | ○ |

| | | | | | | |
|---|---|---|---|---|---|---|
| **Medical management of PrEP/PEP** | ○ | ○ | ○ | ○ | ○ | ○ |
| **Medical management of HIV/AIDS** | ○ | ○ | ○ | ○ | ○ | ○ |

**Notes**: We constructed this question using the answer options of **Question 8** and the format of **Question 6**. This question is therefore a hybrid of questions from Morrison et al.[2] and Obedin-Maliver et al.[1] and original content.

### Survey Part 4: Barriers to Implementation and Strategies for Success

**Question 10: What barriers have you encountered to integrating LGBTQ-related topics into your residency program.**

| | |
|---|---|
| ☐ | Lack of interested faculty |
| ☐ | **Lack of faculty with requisite expertise/knowledge** |
| ☐ | Lack of funding |
| ☐ | Lack of time |
| ☐ | Perception education is not needed |
| ☐ | Opposition to inclusion of LGBTQ content in residency program |
| ☐ | **Influence of state or federal policies and/or political environment** |
| ☐ | **Lack of patient population** |
| ☐ | No barriers |
| ☐ | Other [free form text box] |

**Notes:** This question was adapted from Moll et al. We added a question on faculty expertise to delineate between a lack of interested faculty and a lack of qualified faculty as a barrier to the inclusion of LGBTQ health optics in graduate medical education. We added influence of state/federal policies as an option to reflect recent legislative efforts to limit gender-affirming care in certain states. We added lack of patient population as an answer option to capture a possible sentiment that LGBTQ individuals do not live in certain communities.

**Question 11: What strategies do you think have been or would be successful in increasing LGBTQ-related content at your institution.**

| | **Has been successful in increasing LGBTQ-related content** | **Would likely be successful in increasing LGBTQ-related content** | Don’t know |
|---|---|---|---|
| Curricular material focusing on LGBTQ-related health/health disparities | ○ | ○ | ○ |

| | | | |
|---|---|---|---|
| Faculty willing and able to teach LGBTQ-related content | ◯ | ◯ | ◯ |
| Increased financial resources | ◯ | ◯ | ◯ |
| Logistical support for teaching LGBTQ-related content | ◯ | ◯ | ◯ |
| More time in the curriculum to be able to teach LGBTQ-related content | ◯ | ◯ | ◯ |
| More evidence-based research regarding LGBTQ health/health disparities | ◯ | ◯ | ◯ |
| **More evidence-based research regarding the teaching of LGBTQ-related health topics** | ◯ | ◯ | ◯ |
| Curricular material coverage required by accreditation bodies | ◯ | ◯ | ◯ |
| Questions based on LGBTQ health/health disparities on board examinations | ◯ | ◯ | ◯ |
| Methods to evaluate LGBTQ curricular content | ◯ | ◯ | ◯ |

**Notes:** We adapted this question from Obedin-Maliver et al.[1] We updated the answers to delineate between strategies that programs have utilized to increase LGBTQ content versus strategies programs have not utilized but think would be helpful in increasing LGBTQ content. We added one option on research regarding the teaching of LGBTQ-related health topics as this is (1) 1 burgeoning area of the literature and (2) an area where best practices have not yet been established.

**Supplementary Table 1:** Average Number of Didactic Hours Dedicated to LGBTQ Topics in Graduate Medical Education by Medical Specialty, Stratified by Region and Program Type

| | **Region** | | | | | **Program Type** | | | | |
|---|---|---|---|---|---|---|---|---|---|---|
| | Midwest | Northeast | South | West | P-Value[a] | Community-based | Community-based university affiliated | University-based | Other | P-Value[a] |
| **Specialty (n)** | | | | | | | | | | |
| Anesthesiology (48) | 3.1 | 2.8 | 1.6 | 2.2 | 0.215 | 0.5 | 2.7 | 0.0 | 2.7 | 0.037 |
| Child Neurology (12) | 0.0 | 0.0 | 1.8 | 1.8 | 0.125 | – | 1.5 | – | 1.3 | 0.733 |
| Dermatology (18) | 2.2 | 2.7 | 4.3 | 7.0 | 0.204 | 12.0 | 2.0 | 3.0 | 3.6 | 0.229 |
| Radiology (35) | 2.0 | 1.8 | 2.0 | 1.2 | 0.707 | 2.0 | 1.2 | 12.0 | 1.6 | 0.294 |
| Emergency Medicine (71) | 3.4 | 3.2 | 3.5 | 2.7 | 0.649 | 2.6 | 3.3 | 4.0 | 3.6 | 0.174 |
| Family Medicine (120) | 5.6 | 5.4 | 6.4 | 14.0 | 0.357 | 5.0 | 8.2 | – | 11.0 | 0.003 |
| Internal Medicine (84) | 4.6 | 13.3 | 4.5 | 5.3 | 0.599 | 3.4 | 7.2 | 6.5 | 6.3 | 0.490 |
| Interventional Radiology (7) | 0.0 | 2.0 | 0.3 | – | 0.368 | 0.0 | – | – | 1.2 | 0.408 |
| Neurosurgery (20) | 1.5 | 0.5 | 0.8 | 3.2 | 0.412 | – | 0.8 | – | 1.5 | 0.839 |
| Neurology (44) | 1.6 | 2.7 | 1.7 | 3.9 | 0.396 | 3.3 | 2.7 | 0.0 | 2.2 | 0.225 |
| OBGYN (59) | 4.8 | 4.6 | 4.1 | 6.3 | 0.581 | 4.2 | 4.8 | 4.0 | 4.8 | 0.877 |
| Orthopedic Surgery (27) | 1.3 | 1.7 | 0.6 | 1.5 | 0.730 | 3.0 | 0.8 | – | 1.2 | 0.162 |
| Otolaryngology (18) | 2.9 | 2.5 | 2.3 | 3.7 | 0.944 | 0.0 | 5.0 | – | 2.8 | 0.216 |
| Pathology (23) | 0.8 | 1.2 | 0.9 | 1.0 | 0.993 | – | 0.5 | – | 1.0 | 0.662 |
| Pediatrics (61) | 11.1 | 4.9 | 4.2 | 5.3 | 0.857 | 8.0 | 4.6 | – | 7.1 | 0.392 |
| PM&R (23) | 0.9 | 2.5 | 1.3 | 0.2 | 0.287 | 8.0 | 0.8 | 0.5 | 0.9 | 0.375 |
| Plastic Surgery (6) | 3.5 | 3.0 | 9.0 | 2.0 | 0.456 | – | 2.0 | – | 5.6 | 0.235 |
| Psychiatry (55) | 3.4 | 6.3 | 3.3 | 3.7 | 0.287 | 3.9 | 6.2 | – | 4.3 | 0.576 |
| Public Health (9) | 6.5 | 5.0 | 2.3 | 2.0 | 0.368 | – | 4.0 | – | 3.7 | 0.692 |
| Radiation Oncology (12) | 0.0 | 2.2 | 1.3 | 1.5 | 0.537 | 2.0 | 4.0 | 1.0 | 1.1 | 0.179 |
| Surgery (54) | 1.9 | 3.4 | 1.9 | 1.8 | 0.115 | 0.8 | 2.7 | 2.0 | 2.5 | 0.121 |
| Transition Year (24) | 7.5 | 4.8 | 2.8 | 2.7 | 0.527 | 8.0 | 2.7 | – | 1.2 | 0.077 |
| Urology (23) | 3.3 | 2.0 | 3.2 | 3.7 | 0.382 | 2.0 | 3.2 | – | 2.9 | 0.920 |
| Vascular Surgery (8) | 0.0 | 1.0 | 1.0 | 2.0 | 0.244 | 1.0 | 1.5 | – | 0.6 | 0.409 |
| Other (10) | 4.0 | 0.0 | 5.0 | 0.6 | 0.150 | – | 2.5 | – | 2.1 | 0.343 |

[a]P-Values are the result of the Kruskal-Wallis test.

**Supplementary Table *2*:** Proportion of Programs Dedicating 0 Hours Versus >0 Hours to LGBTQ Didactic and Clinical Content, Stratified by Specialty, Region, and Program Type

| | Didactic Hours | | | Clinical Hours | | |
|---|---|---|---|---|---|---|
| | 0 Hours N (%) | >0 Hours N (%) | P-Value | 0 Hours N (%) | >0 Hours N (%) | P-Value |
| **Specialty** | | | | | | |
| Anesthesiology | 13 (27.1) [15.7–42.1] | 35 (72.9) [57.9–84.2] | < 0.001[a] | 3 (12.0) [3.2–32.3] | 22 (88.0) [67.7–96.8] | < 0.001[a] |
| Child Neurology | 4 (33.3) [11.3–64.6] | 8 (66.7) [35.4–88.7] | | 2 (33.3) [6.0–75.9] | 4 (66.7) [24.1–94.0] | |
| Dermatology | 0 (0.0) [0.0–21.9] | 18 (100.0) [78.1–100.0] | | 2 (25.0) [4.5–64.4] | 6 (75.0) [35.6–95.5] | |
| Diagnostic Radiology | 9 (25.7) [13.1–43.6] | 26 (74.3) [56.4–86.9] | | 13 (68.4) [43.5–86.4] | 6 (31.6) [13.6–56.5] | |
| Emergency Medicine | 1 (1.4) [0.1–8.7] | 70 (98.6) [91.3–100.0] | | 4 (15.4) [5.0–35.7] | 22 (84.6) [64.3–95.0] | |
| Family Medicine | 5 (4.2) [1.5–9.9] | 115 (95.8) [90.0–98.5] | | 7 (7.7) [3.4–15.6] | 84 (91.3) [84.3–96.6] | |
| Internal Medicine | 6 (7.1) [2.9–15.5] | 78 (92.9) [84.5–97.1] | | 7 (12.1) [5.4–23.9] | 51 (87.9) [76.1–94.6] | |
| Interventional Radiology | 4 (57.1) [20.2–88.2] | 3 (42.9) [11.8–79.8] | | – | – | |
| Neurological Surgery | 10 (50.0) [30.0–70.1] | 10 (50.0) [29.9–70.1] | | 4 (36.4) [12.4–68.4] | 7 (63.6) [31.6–87.6] | |
| Neurology | 9 (20.4) [10.3–35.7] | 35 (79.6) [64.2–89.7] | | 5 (22.7) [8.9–45.8] | 17 (77.3) [54.2–91.3] | |
| OBGYN | 2 (3.4) [0.6–12.7] | 57 (96.6) [87.3–99.4] | | 4 (9.5) [3.1–23.5] | 38 (90.5) [76.5–96.9] | |
| Orthopedic Surgery | 12 (44.4) [26.0–64.4] | 15 (55.6) [35.6–74.0] | | 6 (40.0) [17.5–67.1] | 9 (60.0) [32.9–82.5] | |
| Otolaryngology | 5 (27.8) [10.7–53.6] | 13 (72.2) [46.4–89.3] | | 0 (0.0) [0.0–28.3] | 13 (100.0) [71.7–100.0] | |
| Pathology | 11 (47.8) [27.4–68.9] | 12 (52.2) [31.1–72.6] | | 14 (77.8) [52.0–92.6] | 4 (22.2) [7.3–48.1] | |
| Pediatrics | 3 (4.9) [1.3–14.6] | 58 (95.1) [85.4–98.7] | | 1 (2.4) [0.1–14.4] | 40 (97.6) [85.6–99.9] | |
| PM&R | 10 (43.5) [23.9–65.1] | 13 (56.5) [34.9–76.1] | | 7 (53.8) [26.1–79.6] | 6 (46.2) [20.4–73.9] | |
| Plastic Surgery | 0 (0.0) [0.0–48.3] | 6 (100.0) [51.7–100.0] | | – | – | |
| Psychiatry | 1 (1.8) [0.1–11.0] | 54 (98.2) [89.0–100.0] | | 3 (9.7) [2.5–26.9] | 28 (90.3) [73.1–97.5] | |
| Public Health | 2 (22.2) [3.9–59.8] | 7 (77.8) [40.2–96.1] | | 1 (20.0) [1.1–70.1] | 4 (80.0) [29.9–98.9] | |
| Radiation Oncology | 3 (25.0) [6.7–57.2] | 9 (75.0) [42.8–93.3] | | 1 (14.3) [0.8–58.0] | 6 (85.7) [42.0–99.2] | |
| Surgery | 14 (25.9) [15.4–40.0] | 40 (74.1) [60.1–84.6] | | 7 (26.9) [12.4–48.1] | 19 (73.1) [51.9–87.6] | |
| Transition Year | 2 (5.0) [0.8–17.4] | 40 (95.0) [82.6–99.2] | | 3 (25.0) [6.7–57.2] | 9 (75.0) [42.8–93.3] | |
| Urology | 3 (13.0) [3.4–34.7] | 20 (87.0) [65.3–96.6] | | 2 (10.5) [1.8–34.5] | 17 (89.5) [65.5–98.2] | |
| Vascular Surgery | 3 (37.5) [10.2–74.1] | 5 (62.5) [25.9–89.8] | | – | – | |
| Other | 4 (40.0) [13.7–72.6] | 6 (60.0) [27.4–86.3] | | 6 (31.6) [13.6–56.5] | 13 (68.4) [43.5–86.4] | |
| **Region** | | | | | | |
| Midwest | 32 (15.1) [10.7–20.9] | 179 (84.9) [79.1–89.3] | 0.260 | 19 (14.3) [9.0–21.7] | 114 (85.7) [78.3–91.0] | 0.319 |
| Northeast | 29 (12.9) [9.0–18.2] | 195 (87.1) [81.8–91.0] | | 26 (20.0) [13.7–28.1] | 104 (80.0) [71.9–86.3] | |
| South | 53 (19.3) [14.9–24.5] | 222 (80.7) [75.5–85.1] | | 36 (22.9) [16.8–30.4] | 121 (77.1) [69.6–83.2] | |
| West | 24 (14.9) [10.0–21.6] | 137 (85.1) [78.4–90.0] | | 21 (19.6) [12.8–28.7] | 86 (80.4) [71.3–87.2] | |
| **Program Type (%)** | | | | | | |
| Community-based | 19 (15.1) [9.6–22.8] | 107 (84.9) [77.2–90.4] | 0.023 | 15 (21.4) [12.9–33.2] | 55 (78.6) [66.8–87.1] | 0.061 |
| Community-based university affiliated | 33 (11.0) [7.8–15.3] | 266 (89.0) [84.7–92.2] | | 28 (13.6) [9.4–19.2] | 178 (86.4) [80.8–90.6] | |
| Military-based | – | – | | – | – | |
| University-based | 83 (19.1) [15.6–23.2] | 351 (80.9) [76.8–84.4] | | 58 (23.6) [18.5–29.5] | 188 (76.4) [70.5–81.5] | |
| Other | 3 (25.0) [6.7–57.2] | 9 (75.0) [42.8–93.3] | | 1 (20.0) [1.1–70.1] | 4 (80.0) [29.9–98.9] | |

[a]P-values are the result of the Fisher exact test. All other p-values are the result of the Chi-square test.

**Supplementary Table 3:** Average Number of Clinical Hours Dedicated to LGBTQ Patient Care in Graduate Medical Education by Medical Specialty, Stratified by Region and Program Type

| | Region | | | | | Program Type | | | | |
|---|---|---|---|---|---|---|---|---|---|---|
| | Midwest | Northeast | South | West | P-Value[a] | Community-based | Community-based university affiliated | University-based | Other | P-Value[a] |
| **Specialty (n)** | | | | | | | | | | |
| Anesthesiology (25) | 25.0 | 392.5 | 49.1 | 202.8 | 0.2550 | 5.2 | 51.0 | 257.8 | 0.0 | 0.042 |
| Child Neurology (6) | – | – | 10.0 | 7.3 | 1.0000 | – | 0.0 | 10.4 | – | 0.228 |
| Dermatology (8) | 6.7 | 12.0 | 53.3 | 100.0 | 0.3660 | 100.0 | 3.3 | 45.5 | – | 0.079 |
| Radiology (19) | 4.7 | 1.9 | 0.1 | 0.0 | 0.1870 | 5.0 | 1.5 | 0.9 | – | 0.400 |
| Emergency Medicine (26) | 4.0 | 25.2 | 56.9 | 0.7 | 0.0768 | 28.4 | 45.2 | 10.5 | – | 0.386 |
| Family Medicine (91) | 117.0 | 27.6 | 68.8 | 62.0 | 0.3210 | 77.2 | 74.6 | 58.6 | – | 0.225 |
| Internal Medicine (58) | 14.6 | 27.2 | 29.2 | 109.9 | 0.2850 | 53.1 | 28.5 | 61.8 | 11.5 | 0.539 |
| Interventional Radiology (3)[b] | – | – | – | – | – | – | – | – | – | – |
| Neurological Surgery (11) | 15.0 | 0.0 | 346.7 | 17.3 | 0.3680 | – | 347.0 | 12.0 | – | 0.142 |
| Neurology (22) | 50.0 | 5.9 | 29.9 | 15.0 | 0.8300 | – | 4.5 | 28.4 | – | 0.413 |
| OBGYN (42) | 37.5 | 31.6 | 30.3 | 676.5 | 0.1680 | 12.5 | 28.9 | 137.4 | 510.0 | 0.046 |
| Orthopedic Surgery (15) | 10.0 | 7.3 | 0.4 | 162.5 | 0.1940 | 50.0 | 4.6 | 70.1 | – | 0.448 |
| Otolaryngology (13) | 26.7 | 55.0 | 38.3 | 3.5 | 0.3560 | 60.0 | 100.0 | 21.1 | – | 0.161 |
| Pathology (18) | 15.7 | 0.0 | 0.7 | 0.0 | 0.6970 | – | 33.3 | 1.0 | – | 0.464 |
| Pediatrics (41) | 60.3 | 18.4 | 25.1 | 41.6 | 0.2200 | – | 42.0 | 34.6 | – | 0.792 |
| PM&R (13) | 67.7 | 1.0 | 1.0 | 1.2 | 0.6650 | 1.0 | 0.0 | 23.7 | – | 0.253 |
| Plastic Surgery (4)[b] | – | – | – | – | – | – | – | – | – | – |
| Psychiatry (31) | 6.0 | 97.3 | 280.5 | 287.3 | 0.2340 | 10.3 | 73.3 | 324.0 | – | 0.037 |
| Public Health (5) | 30.0 | 0.0 | 30.0 | 10.0 | 0.4430 | – | 30.0 | 13.3 | – | 0.374 |
| Radiation Oncology (7) | 30.0 | 10.0 | 2.0 | 2.0 | 0.4290 | 2.0 | 25.0 | 7.8 | – | 0.549 |
| Surgery (26) | 4.3 | 126.7 | 35.6 | 10.2 | 0.1360 | 5.5 | 12.0 | 100.0 | – | 0.957 |
| Transition Year (12) | 25.4 | 100.0 | 20.0 | 30.0 | 0.2450 | 58.0 | 11.2 | 0.0 | – | 0.038 |
| Urology (19) | 84.2 | 34.0 | 20.0 | 230.0 | 0.5900 | – | 108.7 | 69.4 | – | 0.581 |
| Vascular Surgery (2)[b] | – | – | – | – | – | – | – | – | – | – |
| Other (19) | 168.0 | 5.5 | 36.3 | 51.8 | 0.6810 | – | 26.5 | 63.3 | – | 0.683 |

[a]P-Values are the result of the Kruskal-Wallis test.

[b]These specialties were grouped into the 'Other' category due to having < 5 responses

**Supplementary Table 4:** Free Text Responses for Additional Types of Didactic Education

| Specialty | Response |
|---|---|
| Child Neurology | functional neurologic disorders in LGBTQ |
| Dermatology | We are Derm so most of this not relevant but we do laser hair removal for transgender patients, accutane, cosmetics etcs |
| Family Medicine | Anal pap smears |
| Family Medicine | Human Trafficking and LGBTQ+ populations |
| Family Medicine | Disparities in preventative care |
| Family Medicine | our residency is developing an Affirming care multidisciplinary task force to better meet the needs of our LGBTQ+ population. We have identified access as the #1 barrier in our community |
| Family Medicine | Rural access to GAHT and LGBTQ cultural competency |
| Internal Medicine | Communication with LGBTQ people - understanding and using proper terminology |
| Family Medicine | much of the transgender medicine education comes during the Endocrine rotation which is scheduled during PGY1 year and not all interns are exposed to transgender patients |
| Family Medicine | language / terminology / pronouns |
| Neurosurgery | HIV-related neurological diseases |
| Neurology | There has been no formal information except for HIV/AIDs related infections or end-stage HIV/AIDs |
| Neurology | Pronoun use and Identifying unconscious bias |
| OBGYN | post op care for vaginoplasty patients |
| Orthopedic Surgery | The truth is that I do not know what, if any, topics are formally covered. I think that answer is "none" and I appreciate your study shining a line on this neglect. |
| Orthopedic Surgery | Do not teach this |
| Orthopedic Surgery | Allyship, and musculoskeletal health |
| Pathology | We have discussed how labs should consider reference ranges that may be based on gender, sex, or hormone status |
| Pediatrics | Our residents get bystander and allyship training for the LGBTQ+ population. |
| Pediatrics | Online training required annually for all residents, 1 hour each: Unconscious Bias in the Workplace/Harrassment Prevention: Promoting a Postive Working Environment/REaL and SOGI Cultural Competency Training Program. Sexuality and Healthy Relationships presented by PGY-2 and PGY-3 residents to 500 high school juniors annually. |
| Pediatrics/Medical Genetics | How to provide sensitive genetic/reproductive counseling for LGBTQ people with genetic risk |
| Psychiatry | LGBTQ veterans & Trans population in the correctional system |
| Psychiatry | Interviewing essentials, use of pronouns, appropriate language, WPath |
| Psychiatry | Writing letters for gender-affirming care (we are a psychiatry program) |
| Surgery | Gender affirming surgery and care for this patient population |
| Urology | We have a transgender surgical program |

**Supplementary Table 5:** Free Text Responses for Additional Types of Clinical Education

| Specialty | Response |
|---|---|
| Family Medicine | implicit bias affecting clinical care |
| Family Medicine | LGBTQ primary care |
| Family Medicine | Human Trafficking |
| Family Medicine | LGBTQ dedicated primary care |
| Family Medicine | General medical care, preventative care |
| Family Medicine | primary care of LGBTQ+ and gender diverse patients; pre/post surgical care as PCPs; behavioral health and psychosocial support as PCPs |
| Family Medicine | mental health needs of LGBTQ community |
| Family Medicine | Mental health for LGBTQ+ patients |
| Internal Medicine | Inpatient HIV service. PrEP clinics as a part of primary care clinics. Gender clinic optional. |
| OBGYN | clinical care of LGBTQ not related to their LGBTQ status |
| OBGYN | contraception considerations, safe sexual practices, interpersonal safety, prenatal care for the gender non-conforming/diverse patient |
| OBGYN | health care maintenance |
| OBGYN | Basic health care services and care for birthing individuals |
| OBGYN | None as we are affiliated with a catholic institution that does not allow gender confirming medical or surgical care |
| Otolaryngology | Gender Affirming Voice Therapy (nonsurgical, with SLPs) |
| Otolaryngology | Saw thyroid cartilage shave once as an intern and not since |
| Otolaryngology | gender-affirming voice therapy |
| Otolaryngology | Voice therapy |
| Pediatrics | Affirming care in pediatrics/ adolescents |
| Pediatrics | sexual practices of LGBTQ patients |
| Pediatrics | Mental health management of LGBTQ and gender diverse youth |
| Pediatrics | Mental Health Support |
| Pediatrics | transgender care (all aspects) for children |
| Pediatrics | clinical care of LGBTQ patients in clinic-including STI prevention/treatment, mental health, safe sex practices, substance use, etc |
| Pediatrics | Consent laws for minors regarding hormone therapy. |
| Pediatrics | Behavioral health |
| Pediatrics | mental health, including support letters |
| Urology | orchiectomy |
| Urology | treatment of urologic malignancies and impact |
| Urology | We have a full transgender program - multidisciplinary |

**Supplementary Table 6:** Proportion of Programs Citing Barriers to Incorporating LGBTQ-Content into Graduate Medical Education, Stratified by Specialty, Region, and Program Type

| | Interested Faculty n (%) [CI] | P-Value[a] | Faculty Knowledge n (%) [CI] | P-Value[a] | Funding n (%) [CI] | P-Value[a] |
|---|---|---|---|---|---|---|
| **Specialty (N)** | | | | | | |
| Anesthesiology (39) | 8 (20.5) [9.9–36.9] | 0.415 | 20 (51.3) [35.0–67.3] | 0.061 | 5 (12.8) [4.8–28.2] | 0.258 |
| Child Neurology (8) | 0 (0.0) [0.0–40.2] | | 5 (62.5) [25.9–89.8] | | 0 (0.0) [0.0–40.2] | |
| Dermatology (9) | 2 (22.2) [39.5–59.8] | | 6 (66.7) [30.9–91.0] | | 2 (22.2) [3.9–59.8] | |
| Diagnostic Radiology (22) | 5 (22.7) [8.7–45.8] | | 9 (40.9) [21.5–63.3] | | 2 (9.1) [1.6–30.1] | |
| Emergency Medicine (60) | 11 (18.3) [9.9–30.1] | | 36 (60.0) [46.5–72.2] | | 6 (10.0) [4.1–21.2] | |
| Family Medicine (98) | 22 (22.4) [14.9–32.2] | | 59 (60.2) [50.0–69.8] | | 16 (16.3) [9.9–25.5] | |
| Internal Medicine (67) | 21 (31.3) [20.9–44.0] | | 47 (70.1) [57.6–80.4] | | 14 (20.9) [12.3–32.9] | |
| Interventional Radiology (4)[b] | – | | – | | – | |
| Neurological Surgery (18) | 3 (16.7) [4.4–42.3] | | 9 (50.0) [29.0–71.0] | | 3 (16.7) [4.4–42.3] | |
| Neurology (32) | 4 (12.5) [4.1–29.9] | | 17 (53.1) [35.0–70.5] | | 3 (10.3) [2.7–28.5] | |
| OGBYN (46) | 12 (26.1) [14.8–41.4] | | 30 (65.2) [49.7–78.2] | | 8 (17.4) [8.3–32.0] | |
| Orthopedic Surgery (21) | 6 (28.6) [12.2–52.3] | | 7 (33.3) [15.5–56.9] | | 1 (4.8) [0.2–25.9] | |
| Otolaryngology (15) | 2 (13.3) [2.3–41.6] | | 6 (40.0) [17.5–67.1] | | 1 (6.7) [0.3–34.0] | |
| Pathology (18) | 1 (5.6) [0.3–29.4] | | 8 (44.4) [22.4–68.7] | | 1 (5.6) [0.3–29.4] | |
| Pediatrics (45) | 4 (8.9) [2.9–22.1] | | 25 (53.3) [38.0–68.1] | | 12 (26.7) [15.1–42.2] | |
| PM&R (19) | 4 (21.1) [7.0–46.1] | | 13 (68.4) [43.5–86.4] | | 0 (0.0) [0.0–20.9] | |
| Plastic Surgery (4)[b] | – | | – | | – | |
| Psychiatry (38) | 5 (13.2) [4.9–28.9] | | 21 (55.3) [38.5–71.0] | | 7 (18.4) [8.3–34.9] | |
| Public Health (5) | 0 (0.0) [0.0–53.7] | | 3 (60.0) [17.0–92.7] | | 2 (40.0) [7.3–83.0] | |
| Radiation Oncology (8) | 0 (0.0) [0.0–40.2] | | 3 (37.5) [10.2–74.1] | | 0 (0.0) [0.0–40.2] | |
| Surgery (35) | 8 (22.9) [11.0–40.6] | | 21 (60.0) [42.2–75.6] | | 6 (17.1) [7.2–34.3] | |
| Transition Year (13) | 2 (18.2) [3.2–52.2] | | 10 (76.9) [46.0–93.8] | | 4 (44.4) [15.3–77.3] | |
| Urology (17) | 5 (29.4) [11.4–56.0] | | 6 (35.3) [15.3–61.4] | | 2 (11.8) [2.1–37.7] | |
| Vascular Surgery (5) | 1 (25.0) [1.3–78.1] | | 3 (60.0) [17.0–92.7] | | 1 (25.0) [1.3–78.1] | |
| Other (18) | 3 (16.7) [4.4–42.3] | | 5 (27.7) [10.7–53.6] | | 2 (11.1) [1.9–36.1] | |
| **Region (N)** | | | | | | |
| Midwest | 39 (23.2) [17.2–30.5] | 0.519 | 103 (61.3) [53.5–68.6] | 0.026 | 28 (16.7) [11.5–23.4] | 0.829 |
| Northeast | 29 (18.4) [12.8–25.5] | | 84 (53.2) [45.1–61.1] | | 22 (14.0) [9.1–20.5] | |
| South | 39 (19.7) [14.5–26.0] | | 120 (60.1) [53.4–67.4] | | 27 (13.6) [9.3–19.4] | |
| West | 22 (16.7) [11.0–24.4] | | 61 (46.2) [37.6–55.1] | | 21 (15.9) [10.3–23.5] | |
| **Program Type (N)** | | | | | | |
| Community-based | 27 (28.7) [20.1–39.1] | 0.002 | 55 (58.5) [47.9–68.4] | 0.001 | 15 (16.0) [9.6–25.3] | 0.802 |
| Community-based university-affiliated | 50 (21.8) [16.8–27.9] | | 147 (64.2) [57.6–70.3] | | 39 (17.0) [12.5–22.7] | |
| University-based | 49 (15.1) [11.5–19.5] | | 162 (49.9) [44.3–55.4] | | 42 (12.9) [9.6–17.2] | |
| Other | 3 (37.5) [10.2–74.1] | | 4 (50.0) [21.5–78.5] | | 2 (25.0) [4.5–64.4] | |

| | Lack of Time N (%) [CI] | P-Value | Perception Education is Not Needed N (%) [CI] | P-Value | Opposition N (%) [CI] | P-Value |
|---|---|---|---|---|---|---|
| **Specialty** | | | | | | |
| Anesthesiology (39) | 17 (43.6) [28.2–60.2] | 0.058 | 15 (38.5) [23.8–55.3] | < 0.001 | 3 (7.7) [2.0–22.0] | 0.906 |
| Child Neurology (8) | 4 (50.0) [21.5–78.5] | | 0 (0.0) [0.0–40.2] | | 0 (0.0) [0.0–40.2] | |
| Dermatology (9) | 6 (66.7) [30.9–91.0] | | 1 (11.1) [0.6–49.3] | | 1 (12.5) [0.7–53.3] | |
| Diagnostic Radiology (22) | 4 (18.2) [6.0–41.0] | | 6 (27.3) [11.6–50.4] | | 0 (0.0) [0.0–18.5] | |
| Emergency Medicine (60) | 37 (61.7) [48.2–73.6] | | 9 (15.0) [7.5–27.1] | | 6 (10.0) [4.1–22.2] | |
| Family Medicine (98) | 57 (58.2) [47.8–67.0] | | 9 (10.1) [5.0–18.8] | | 7 (7.1) [3.2–14.7] | |
| Internal Medicine (67) | 36 (53.7) [41.2–65.8] | | 10 (14.9) [7.8–26.2] | | 4 (6.0) [1.9–15.3] | |
| Interventional Radiology (4)[b] | – | | – | | – | |
| Neurological Surgery (18) | 8 (44.4) [22.4–68.7] | | 9 (50.0) [29.0–71.0] | | 2 (11.1) [1.9–36.1] | |
| Neurology (32) | 17 (53.1) [35.0–70.5] | | 6 (18.8) [7.9–37.0] | | 1 (3.1) [0.2–18.0] | |
| OGBYN (46) | 24 (57.2) [37.1–66.9] | | 2 (4.3) [0.8–16.0] | | 0 (0.0) [0..0–9.6] | |
| Orthopedic Surgery (21) | 7 (33.3) [15.5–56.9] | | 6 (28.6) [12.2–52.3] | | 2 (9.5) [1.7–31.8] | |
| Otolaryngology (15) | 6 (40.0) [17.5–67.1] | | 3 (20.0) [5.3–48.6] | | 1 (6.7) [0.3–34.0] | |
| Pathology (18) | 5 (27.8) [10.7–53.6] | | 4 (22.2) [7.4–48.1] | | 1 (5.6) [0.3–29.4] | |
| Pediatrics (45) | 22 (44.4) [30.0–59.9] | | 2 (4.4) [0.8–16.4] | | 4 (8.9) [2.8–22.1] | |
| PM&R (19) | 8 (72.3) [39.3–92.7] | | 5 (26.3) [10.1–51.4] | | 1 (5.3) [0.3–28.1] | |
| Plastic Surgery (4)[b] | – | | – | | – | |
| Psychiatry (38) | 18 (44.7) [29.0–61.5] | | 2 (5.3) [0.9–19.1] | | 2 (5.3) [0.9–19.1] | |
| Public Health (5) | 3 (60.0) [17.0–92.7] | | 0 (0.0) [0.0–53.7] | | 0 (0.0) [0.0–53.7] | |
| Radiation Oncology (8) | 2 (25.0) [4.5–64.4] | | 3 (37.5) [10.2–74.1] | | 0 (0.0) [0.0–40.2] | |
| Surgery (35) | 20 (57.1) [40.0–73.2] | | 7 (20.0) [9.1–37.5] | | 2 (5.7) [1.0–20.5] | |
| Transition Year (13) | 6 (46.2) [20.4–74.0] | | 0 (0.0) [0.0–28.3] | | 0 (0.0) [0.0–28.3] | |
| Urology (17) | 6 (35.3) [15.3–61.4] | | 3 (17.6) [4.7–44.2] | | 2 (11.8) [2.1–37.7] | |
| Vascular Surgery (5) | 1 (20.0) [1.1–70.1] | | 2 (40.0) [7.3–83.0] | | 0 (0.0) [0.0–53.7] | |
| Other (18) | 6 (33.3) [14.4–58.8] | | 3 (16.7) [4.4–42.3] | | 1 (5.6) [0.3–29.4] | |
| **Region** | | | | | | |
| Midwest | 81 (48.2) [40.5–56.0] | 0.976 | 26 (15.5) [10.5–22.0] | 0.268 | 11 (6.5) [3.5–11.7] | 0.012 |
| Northeast | 76 (48.1) [40.1–56.2] | | 27 (17.1) [11.8–24.1] | | 3 (1.9) [0.5–5.9] | |
| South | 94 (47.5) [40.1–52.2] | | 26 (13.1) [8.9–18.8] | | 20 (11.2) [7.2–17.0] | |
| West | 66 (50.0) [41.6–58.4] | | 28 (21.2) [14.8–29.4] | | 6 (4.5) [1.9–10.1] | |
| **Program Type** | | | | | | |
| Community-based | 38 (40.4) [30.6–51.1] | 0.215 | 14 (14.9) [8.7–24.1] | 0.104 | 8 (8.5) [4.0–16.6] | 0.401 |
| Community-based university-affiliated | 110 (48.0) [41.4–54.7] | | 32 (14.0) [9.9–19.3] | | 9 (3.9) [1.9–7.6] | |
| University-based | 166 (50.0) [44.1–55.0] | | 59 (18.2) [14.2–22.9] | | 23 (7.1) [4.6–10.6] | |
| Other | 3 (37.5) [10.2–74.1] | | 2 (25.0) [4.5–64.4] | | 0 (0.0) [0.0–40.2] | |
| | **State/Federal Policies** | **P-Value** | **Lack of Patients** | **P-Value** | **No Barriers** | **P-Value** |

| | N (%) [CI] | | N (%) [CI] | | N (%) [CI] | |
|---|---|---|---|---|---|---|
| **Specialty** | | | | | | |
| Anesthesiology (39) | 8 (20.5) [9.9–36.9] | | 5 (12.8) [4.8–28.2] | | 10 (25.6) [13.6–42.4] | |
| Child Neurology (8) | 0 (0.0) [0.0–40.2] | | 0 (0.0) [0.0–40.2] | | 3 (25.0) [4.5–64.4] | |
| Dermatology (9) | 2 (22.2) [3.9–59.8] | | 1 (11.1) [0.6–49.3] | | 0 (0.0) [0.0–37.1] | |
| Diagnostic Radiology (22) | 0 (0.0) [0.0–18.5] | | 4 (18.2) [6.0–41.0] | | 14 (63.6) [40.8–82.0] | |
| Emergency Medicine (60) | 10 (16.7) [8.7–29.0] | | 4 (6.7) [2.2–17.0] | | 8 (13.3) [6.3–25.1] | |
| Family Medicine (98) | 19 (19.4) [12.4–28.9] | | 16 (16.3) [9.9–25.5] | | 10 (10.2.) [5.3–18.4] | |
| Internal Medicine (67) | 10 (14.9) [7.8–26.2] | | 12 (17.9) [10.0–29.6] | | 4 (6.0) [1.9–15.3] | |
| Interventional Radiology (4)[b] | – | | – | | – | |
| Neurological Surgery (18) | 1 (5.6) [0.3–29.4] | | 3 (16.7) [4.4–42.3] | | 8 (44.4) [22.4–68.7] | |
| Neurology (32) | 3 (9.4) [2.5–26.2] | | 4 (12.5) [4.1–29.9] | | 8 (25.0) [12.1–43.8] | |
| OGBYN (46) | 5 (10.9) [4.1–24.4] | | 8 (17.4) [8.3–32.0] | | 7 (15.2) [6.8–29.5] | |
| Orthopedic Surgery (21) | 2 (9.5) [1.7–31.8] | | 4 (19.0) [6.3–42.6] | | 8 (38.1) [19.0–61.3] | |
| Otolaryngology (15) | 2 (13.3) [2.3–41.6] | 0.015 | 5 (33.3) [13.0–61.3] | 0.566 | 5 (33.3) [13.0–61.3] | < 0.001 |
| Pathology (18) | 2 (11.1) [1.9–36.1] | | 1 (5.6) [0.3–29.4] | | 6 (33.3) [14.4–58.8] | |
| Pediatrics (45) | 17 (37.8) [24.2–53.5] | | 3 (6.7) [1.7–19.3] | | 10 (22.2) [11.7–37.5] | |
| PM&R (19) | 1 (5.3) [0.3–28.1] | | 3 (15.8) [4.2–40.5] | | 3 (15.8) [4.2–40.5] | |
| Plastic Surgery (4)[b] | – | | – | | – | |
| Psychiatry (38) | 2 (5.3) [0.9–19.1] | | 4 (10.5) [3.4–25.7] | | 9 (23.7) [12.0–40.6] | |
| Public Health (5) | 1 (20.0) [1.1–70.1] | | 0 (0.0) [0.0–53.7] | | 1 (20.0) [1.1–70.1] | |
| Radiation Oncology (8) | 0 (0.0) [0.0–40.2] | | 0 (0.0) [0.0–40.2] | | 1 (12.5) [0.7–53.3] | |
| Surgery (35) | 2 (5.7) [0.1–20.5] | | 3 (8.6) [2.2–24.2] | | 9 (25.7) [13.1–43.6] | |
| Transition Year (13) | 4 (30.8) [10.4–61.1] | | 2 (15.4) [2.7–46.3] | | 2 (15.4) [2.7–46.3] | |
| Urology (17) | 3 (17.6) [4.7–44.2] | | 4 (23.5) [7.8–50.2] | | 5 (29.4) [11.4–56.0] | |
| Vascular Surgery (5) | 0 (0.0) [0.0–53.7] | | 0 (0.0) [0.0–53.7] | | 2 (40.0) [7.3–83.0] | |
| Other (18) | 2 (11.1) [1.9–36.1] | | 4 (22.2) [7.4–48.0] | | 9 (50.0) [29.0–70.9] | |
| **Region** | | | | | | |
| Midwest | 28 (16.7) [11.5–23.4] | | 25 (14.9) [10.0–21.4] | | 33 (19.6) [14.1–26.6] | |
| Northeast | 4 (2.5) [0.8–6.7] | < 0.001 | 25 (15.8) [10.7–22.7] | 0.659 | 36 (22.8) [16.7–30.3] | 0.137 |
| South | 56 (28.3) [22.2–35.2] | | 25 (12.6) [8.5–18.3] | | 35 (17.7) [12.8–23.9] | |
| West | 8 (6.1) [2.8–12.0] | | 15 (11.4) [6.7–18.3] | | 37 (28.0) [20.8–36.6] | |
| **Program Type** | | | | | | |
| Community-based | 16 (17.0) [10.3–26.5] | | 15 (16.0) [9.5–25.3] | | 18 (19.1) [12.0–28.8] | |
| Community-based university-affiliated | 35 (15.3) [11.0–20.8] | 0.928 | 33 (14.4) [10.3–19.8] | 0.089 | 45 (19.7) [14.8–25.5] | 0.079 |
| University-based | 45 (13.8) [10.4–18.2 | | 42 (12.9) [9.6–17.2] | | 75 (30.0) [24.5–36.2] | |
| Other | 0 (0.0) [0.0–40.2] | | 0 (0.0) [0.0–40.2] | | 3 (37.5) [10.2–74.1 | |

[a]P-values for specialty are the result of the Fisher exact test. All other p-values are the result of the Chi-square test.
[b]Interventional radiology and plastic surgery are grouped into the ‘Other’ category since they have < 5 responses
